# Excess deaths in Spain during the first year of the COVID–19 pandemic outbreak from age/sex–adjusted death rates

**DOI:** 10.1101/2020.07.22.20159707

**Authors:** José María Martín-Olalla

## Abstract

**Objectives:** Assess the impact of the illness designated COVID–19 during the first year of pandemic outbreak in Spain through age/sex–specific death rates.

**Study design:** Age/sex–specific weeekly deaths in Spain were retrieved from Eurostat. Spanish resident population was obtained from the National Statistics Office.

**Methods:** Generalized linear Poisson regressions were used to compute the contrafactual expected rates after one year (52 weeks or 364 days) of the pandemic onset. From this one–year age/sex–specific and age/sex–adjusted mortality excess rates were deduced.

**Results:** For the past continued 13 years one–year age/sex–adjusted death rates had not been as high as the rate observed on February 28th, 2021.

The excess death rate was estimated as 1.790×10^−3^ (95 % confidence interval, 1.773×10^−3^ to 1.808×10^−3^; *P*−score = 20.2 % and *z*−score = 11.4) with an unbiased standard deviation of the residuals equal to 157×10^−6^. This made 84 849 excess deaths (84 008 to 85 690). Sex disaggregation resulted in 44 887 (44 470 to 45 303) male excess deaths and 39 947 (39 524 to 40 371) female excess deaths.

**Conclusion:** With 73 571 COVID–19 deaths and 9772 COVID–19 suspected deaths that occurred in nursing homes during the spring of 2020 it is only 1496 excess deaths (1.8 %, a *z*−score of 0.2) that remains unattributed.

The infection rate during the first year of the pandemic is estimated in 16 % of population after comparing the ENE–COVID seroprevalence, the excess deaths at the end of the spring 2020 and the excess deaths at the end of the first year of the pandemic.

## 1. INTRODUCTION

The illness designated covid–19 caused by the severe acute respiratory syndrome coronavirus 2 (SARS–CoV–2) caught worldwide attention since its identification in late 2019[8]. Spain was one of the European countries most impacted by the disease during the spring of 2020[3]. Confirmed covid–19 cases climbed up to several hundred thousands —some few thousands per one million population— and confirmed covid–19 deaths to some 70 000 or 1500 deaths per one million population. A myriad of societal measures ranging from a lock–down to a mandatory use of masks were enforced since the outbreak of the pandemic.

Total excess deaths or crude excess deaths or all–cause excess deaths is a key quantity to understand the impact of a pandemic[1]. Many efforts and resources are dedicated to provided fast and accurate numbers of this societal quantity (see Islam *et al*. [4] (OECD countries) and Karlinsky and Kobak [5] (world wide)). While the observed deaths during a window of time can be accurately known from official records in modern societies the determination of a predicted or reference value from which excesses are computed is a matter of retrospective statistical analysis. Mortality is characteristically different for males and females and for age groups. Therefore the understanding of age–specific, sex–specific death rates is of the utmost importance. Their trends highlight societal issues: either improvement of public health care which leads to smaller specific rates or, events like wars, outbreaks or economic turmoils which lead to greater specific rates. Age/sex–specific mortality rates are the basic quantities to assess statistical measures like the life expectancy which are also object of interest in this context[7].

In this paper I analyse Spanish official records of weekly mortality and population dated back from the year 2000. The manuscript is devoted the accumulated mortality in the first 52 weeks (364 days or one year) after the pandemic onset. Total excesses will be derived for averaged and adjusted age/sex specific death rates for the last available structure of the Spanish population. Infection rate during the first year of the pandemic will be derived from SARS–CoV–2 seroprevalence.

## 2. METHODS

### 2.1. Data sets

Eurostat file `demo_r_mweek3.tsv` lists weekly deaths provided National Statistics Office since, at most, the year 2000. Data are disaggregated by statistical region (all levels), sex (males, females, both), and age group (total, five–year age groups until the group 90 years old and above). Weeks follow the ISO week date convention. The file was downloaded on August 11th, 2021 and includes the last major revision by INE of the 2020 weekly deaths, released on June 17th, 2021. However, it should be noted that 2020 and 2021 weekly deaths are still flagged as “provisional”. The Instituto Nacional de Estadística (INE) table `31304` lists resident population values in Spain for January 1st and July 1st since the year 1971 until January 1st, 2021. The data set is disaggregated by statistical region (NUTS3), sex and age until the group 85 years old and above.

Two steps were taken in order to harmonised both sets. First, weekly deaths for the age group 85–89 and the age group 90 or elder were summed up to yield the age group 85 or elder. Second, population numbers for January 1st and July 1st were assigned to their corresponding week dates. Then population numbers for every week since 2000 were computed by interpolation. The last available numbers (July 1st, 2020) were extended into 2021 for the lack of a better quantity.

With these, 52–week, all–cause deaths were computed. This quantity accumulates the deaths observed on a given week and on the preceding 51 weeks, for a total of 364 days or one year in the context of this manuscript. The 52–week accumulated values were then scaled by the population at the given week to get the 52–week accumulated death rate.

In addition to that confirmed COVID–19 cases and deaths are numbered in Spain by the Centro Nacional de Epidemiología (CNE), equivalent to the CDC after reports from local authorities. The data set is disaggregated by sex, NUTS3 region and 10–year age groups on a daily basis. These records were harmonised to read weekly counts.

### 2.2. Statistical analysis

The pre–pandemic period in Spain ended on Sunday March 1st, 2020. One year (364 days) later it came Sunday February 28th 2021 at the end of week 08–2021.

To account for the seasonality of human mortality 52–week accumulated death rates at week 08 on every year since 2001 were computed from the data set. A bivariate generalised Poisson regression with a linear rate served to infer the predicted 52–week death rate for the week 08–2021. The predictors of the regression were the ordinal week numbers. The responses were the observed 52–week death rates. The regression was restricted to the eight years prior the pandemic onset: from 2013 to 2020. The unbiased standard deviation of the residuals was recorded.

The bivariate regression was performed for age–specific, sex–specific death rates *d*_*ij*_ (*i* refers to sex; *j*, to age group). The analysis was complemented with two adjusted analysis. First the regression of the sex–specific, age–adjusted weekly death rate defined as:

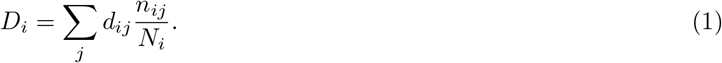

where *n*_*ij*_ are the age–specific, sex–specific last population numbers in Spain and *N*_*i*_ the total number of persons with sex *i*. And second, the regression of the age/sex–adjusted weekly death rate, defined as :

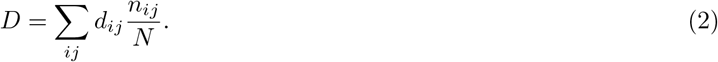

where *N* is the Spanish population.

Notice that in Eq. (1) and in Eq. (2) the rate *d*_*ij*_ depends on the week date, therefore they refer to different years across the catalogue. However the population numbers *n*_*ij*_ and *N* are those given by the last available records of population. Therefore adjusted weekly death rates propagate the population pyramid back and forth in time so that *D* or *D*_*i*_ only track the changes in the specific mortality. Compared to that, the unspecific death rates —total deaths scaled by total population— track changes in specific mortality and in population composition.

Usually in age–harmonised analysis the weights are given by a standard, invariant population pyramid, often distant in time. As an example the European Standard Population[2] is widely used to facilitate the comparison between countries, see for instance Islam *et al*. [4]. However since the purpose of this manuscript is the comparison of the Spanish mortality throughout the 21st century I opted to extend the population structure in 2021 back in time to compute the death rate that would have been observed in Spain during 2021 if the age/sex–specific death rates for 2021 had been those observed in the preceding years. In this way excesses deduced from this age/sex–adjusted rates are meaningful in 2021.

## 3. RESULTS

Figure 1 shows the age–specific, sex–specific, all–cause, 52–week death rate in Spain since the year 2001. Blueish lines and circles refer to the male distribution; orange lines and squares refer to the female distribution. Notice that sex–unspecific sample average and sample standard deviation were removed from the data set so that the vertical axes span equally through different age groups even though observed death rates are vividly different.

**FIG. 1.**
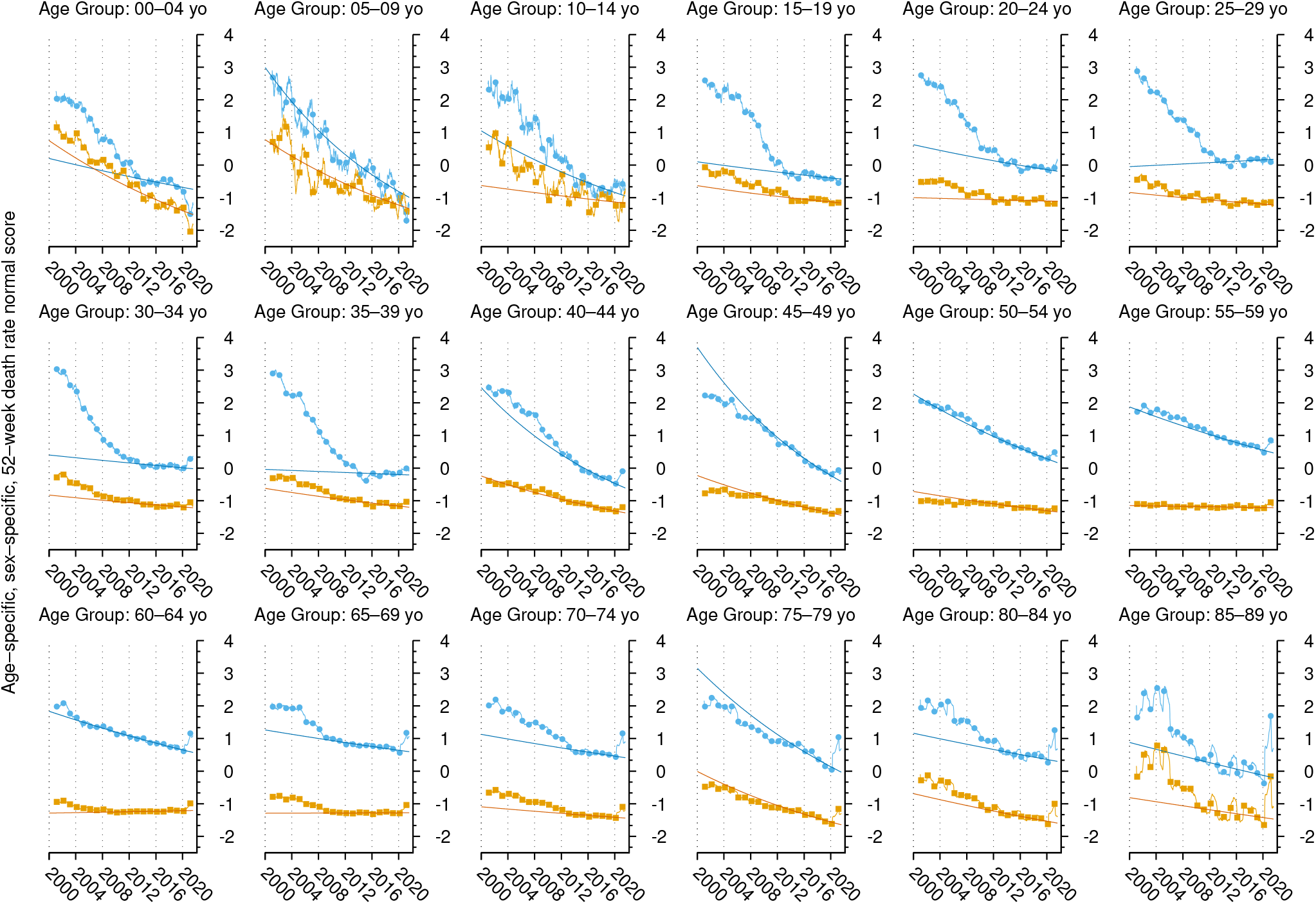
Age/sex–specific 52–week death rate normal scores. Data points show values at week 08 on every year since 2001 (blue circles, males; orange squares, females). They are joined by weekly values in the remaining weeks of the year (thin light line). Solid dark line displays the results of the Poisson regression for the eight years prior to the pandemic. The normal scores were computed as (*x*−*E*)*/s* where *E* is the sample average and *s* the sample standard deviation of the age–specific, sex–unspecific data set.

Death rates in Figure 1 show a decreasing trend with increasing year as a result of enhancement in societal and public health issues. Some age groups —notably the younger age groups, but also some older groups like the 65–69 age group— show a stalling in the past few years.

Pondering every aspect of this distribution I choose to infer the expected death rate in week 08–2021 from a generalised Poisson regression that included the eight years prior to the pandemic. Regression results are shown in Figure 1 by a darker line often deviating from data points in the first records of the data set. Generally speaking the observed death rate in 2021 shows an excursion for male population elder than 30 and female population elder than 40.

For each stratum, Table 1 lists the 2021 all–cause excess death rate with estimate and 95 % confidence interval, the 2021 all–cause total excess deaths (estimate and 95 % CI), the *P*—score (ratio excess to predicted value), the *z*−score (ratio excess to unbiased standard deviation of the residuals) and the number of years since a 52–week death rate greater than the observed 2021 52–week death rate was last recorded. Finally, the table lists the COVID–19 undercount (ratio of excess deaths to COVID–19 deaths) and the COVID–19 case fatality risks (CFR, ratio of excess deaths to laboratory confirmed COVID–19 cases). Notice that COVID–19 cases were undercount until June 2020 notably in younger age groups, more prone to a mild evolution of the disease.

**TABLE 1.** Age/sex–specific, all–cause mortality data in Spain during the first year of the pandemic. First column lists the estimate and the 95 % confidence interval of the excess death rate, followed by estimate and 95 % CI for total excess deaths. Then the *P*–score (the ratio of the excess to the expected deaths, given as a percent); the *z*−score (the ratio of the excess to the unbiased sample standard deviation of the residuals); and the number of years since the observed death rate was larger than the one observed in the year 2021, with *>* underscoring a period longer than the record size of 20 years. The two columns list the ratio of 52–week excess deaths to COVID–19 52–week deaths (the undercount) and the ratio of 52–week excess deaths to 52–week COVID–19 cases, the case fatality risk CFR, expressed as a percentage. The “all ages” strata refer to the adjusted rates (see Eq. (1) and Eq. (2)).

| Stratum | 2021 all-cause excess death rate<br>estimate [95 % CI]/10<sup>−3</sup> |  |  | 2021 all-cause total excess<br>estimate [95 % CI] |  |  | *P*-score<br>/% | *z*-score | Years<br>back | COVID-19<br>undercount | COVID-19<br>CFR/% |
| --- | --- | --- | --- | --- | --- | --- | --- | --- | --- | --- | --- |
| All sexes |  |  |  |  |  |  |  |  |  |  |  |
| All ages | 1.790 | [ 1.773 to 1.808 ] |  | 84 849 | [ 84 008 to 85 690 ] |  | 20.2 | 11.4 | 12 | 1.153 | 2.73 |
| Males |  |  |  |  |  |  |  |  |  |  |  |
| All ages | 1.933 | [ 1.915 to 1.951 ] |  | 44 887 | [ 44 470 to 45 303 ] |  | 21.2 | 15.2 | 12 | 1.112 | 3.05 |
| 00–04 | −0.142 | [ −0.190 to −0.095 ] |  | −141 | [ −187 to −94 ] |  | −22.5 | −6.5 | 1 |  |  |
| 05–09 | −0.0202 | [ −0.0248 to −0.0156 ] |  | −24 | [ −29 to −18 ] |  | −30.3 | −4.0 | 1 |  |  |
| 10–14 | 0.0111 | [ 0.0059 to 0.0162 ] |  | 14 | [ 8 to 21 ] |  | 14.3 | 1.6 | 6 |  |  |
| 15–19 | −0.0178 | [ −0.0263 to −0.0094 ] |  | −22 | [ −33 to −12 ] |  | −8.8 | −1.8 | 1 |  |  |
| 20–24 | 0.016 | [ 0.006 to 0.027 ] |  | 20 | [ 7 to 33 ] |  | 5.1 | 1.0 | 2 |  |  |
| 25–29 | −0.015 | [ −0.027 to −0.002 ] |  | −19 | [ −35 to −3 ] |  | −3.5 | −0.8 | 1 | 0.023 |  |
| 30–34 | 0.070 | [ 0.057 to 0.084 ] |  | 96 | [ 78 to 115 ] |  | 13.7 | 5.9 | 10 |  |  |
| 35–39 | 0.065 | [ 0.049 to 0.081 ] |  | 103 | [ 78 to 128 ] |  | 9.3 | 1.4 | 8 | 1.782 | 0.10 |
| 40–44 | 0.223 | [ 0.205 to 0.240 ] |  | 434 | [ 400 to 468 ] |  | 23.6 | 6.7 | 6 |  |  |
| 45–49 | 0.205 | [ 0.182 to 0.229 ] |  | 407 | [ 360 to 454 ] |  | 11.7 | 5.9 | 3 | 1.801 | 0.34 |
| 50–54 | 0.315 | [ 0.281 to 0.349 ] |  | 582 | [ 519 to 646 ] |  | 9.2 | 10.0 | 4 |  |  |
| 55–59 | 0.628 | [ 0.583 to 0.672 ] |  | 1060 | [ 984 to 1135 ] |  | 11.0 | 8.3 | 7 | 0.941 | 0.74 |
| 60–64 | 1.520 | [ 1.464 to 1.576 ] |  | 2227 | [ 2144 to 2309 ] |  | 17.0 | 16.0 | 12 |  |  |
| 65–69 | 2.380 | [ 2.309 to 2.450 ] |  | 2826 | [ 2743 to 2910 ] |  | 17.1 | 12.8 | 13 | 1.052 | 3.31 |
| 70–74 | 4.670 | [ 4.583 to 4.756 ] |  | 4786 | [ 4697 to 4875 ] |  | 22.2 | 15.3 | 11 |  |  |
| 75–79 | 9.88 | [ 9.55 to 10.21 ] |  | 7819 | [ 7558 to 8079 ] |  | 31 | 9.4 | 11 | 1.205 | 12.59 |
| 80–84 | 12.70 | [ 12.23 to 13.18 ] |  | 6780 | [ 6525 to 7034 ] |  | 19.9 | 14.6 | 12 |  |  |
| ≥ 85 | 32.94 | [ 32.23 to 33.66 ] |  | 17 773 | [ 17 388 to 18 158 ] |  | 23.2 | 10.1 | 16 | 1.080 | 28.52 |
| Females |  |  |  |  |  |  |  |  |  |  |  |
| All ages | 1.653 | [ 1.635 to 1.670 ] |  | 39 947 | [ 39 524 to 40 371 ] |  | 19.2 | 8.7 | 14 | 1.204 | 2.44 |
| 00–04 | −0.101 | [ −0.142 to −0.060 ] |  | −94 | [ −132 to −56 ] |  | −20.5 | −3.7 | 1 |  |  |
| 05–09 | −0.0031 | [ −0.0075 to 0.0013 ] |  | −3 | [ −8 to 1 ] |  | −5.5 | −0.7 | 1 |  |  |
| 10–14 | −0.0020 | [ −0.0070 to 0.0029 ] |  | −2 | [ −9 to 4 ] |  | −2.9 | −0.2 | 2 |  |  |
| 15–19 | 0.000 | [ −0.0060 to 0.0060 ] |  | 0 | [ −7 to 7 ] |  | 0.0 | 0.0 | 2 |  |  |
| 20–24 | −0.0147 | [ −0.0220 to −0.0075 ] |  | −17 | [ −26 to −9 ] |  | −10.2 | −1.1 | 1 |  |  |
| 25–29 | 0.0146 | [ 0.0070 to 0.0222 ] |  | 18 | [ 9 to 28 ] |  | 8.8 | 0.9 | 4 | 0.029 |  |
| 30–34 | 0.0380 | [ 0.0291 to 0.0469 ] |  | 52 | [ 40 to 64 ] |  | 16.7 | 3.5 | 8 |  |  |
| 35–39 | 0.052 | [ 0.041 to 0.064 ] |  | 84 | [ 66 to 103 ] |  | 14.3 | 2.6 | 7 | 1.796 | 0.06 |
| 40–44 | 0.074 | [ 0.060 to 0.088 ] |  | 144 | [ 117 to 171 ] |  | 12.9 | 6.3 | 5 |  |  |
| 45–49 | 0.075 | [ 0.056 to 0.093 ] |  | 146 | [ 110 to 182 ] |  | 7.3 | 4.2 | 3 | 1.168 | 0.10 |
| 50–54 | 0.110 | [ 0.086 to 0.135 ] |  | 204 | [ 158 to 250 ] |  | 6.3 | 2.9 | 3 |  |  |
| 55–59 | 0.275 | [ 0.243 to 0.307 ] |  | 478 | [ 423 to 534 ] |  | 9.8 | 4.3 | > | 0.884 | 0.28 |
| 60–64 | 0.592 | [ 0.553 to 0.631 ] |  | 919 | [ 859 to 979 ] |  | 14.4 | 12.3 | 19 |  |  |
| 65–69 | 1.005 | [ 0.959 to 1.052 ] |  | 1310 | [ 1249 to 1371 ] |  | 16.8 | 9.9 | 15 | 1.187 | 1.47 |
| 70–74 | 2.139 | [ 2.081 to 2.196 ] |  | 2546 | [ 2477 to 2614 ] |  | 23.0 | 16.5 | 11 |  |  |
| 75–79 | 4.47 | [ 4.24 to 4.71 ] |  | 4429 | [ 4195 to 4663 ] |  | 27 | 8.4 | 9 | 1.323 | 6.68 |
| 80–84 | 7.80 | [ 7.44 to 8.17 ] |  | 6039 | [ 5754 to 6323 ] |  | 20.4 | 11.1 | 12 |  |  |
| ≥ 85 | 22.73 | [ 22.53 to 22.94 ] |  | 23 578 | [ 23 362 to 23 793 ] |  | 19.0 | 6.8 | 16 | 1.190 | 19.81 |

In addition to age/sex–specific analyses, Table 1 also shows the age/sex–adjusted analysis for every sex and for the whole population. In order to provide a better visualisation of the regression method Figure 2 shows the age/sex–adjusted, all–cause weekly death rate (bottom) and the age/sex–adjusted all–cause 52–week cumulative death rate (top, dark orange). For the sake of comparison the unspecific (all–age, all–sex) 52–week death rate is shown in light grey. Notice that the unspecific death rate and the adjusted death rate match to each other at the end of the record as per construction. When they differ the adjusted death rate displays the death rate that would have been observed in the year 2021 had population died at the age–specific, sex–specific death rates observed in the previous weeks/years. The age–unspecific, sex–unspecific death rate displays the actual observed death rate.

**FIG. 2.**
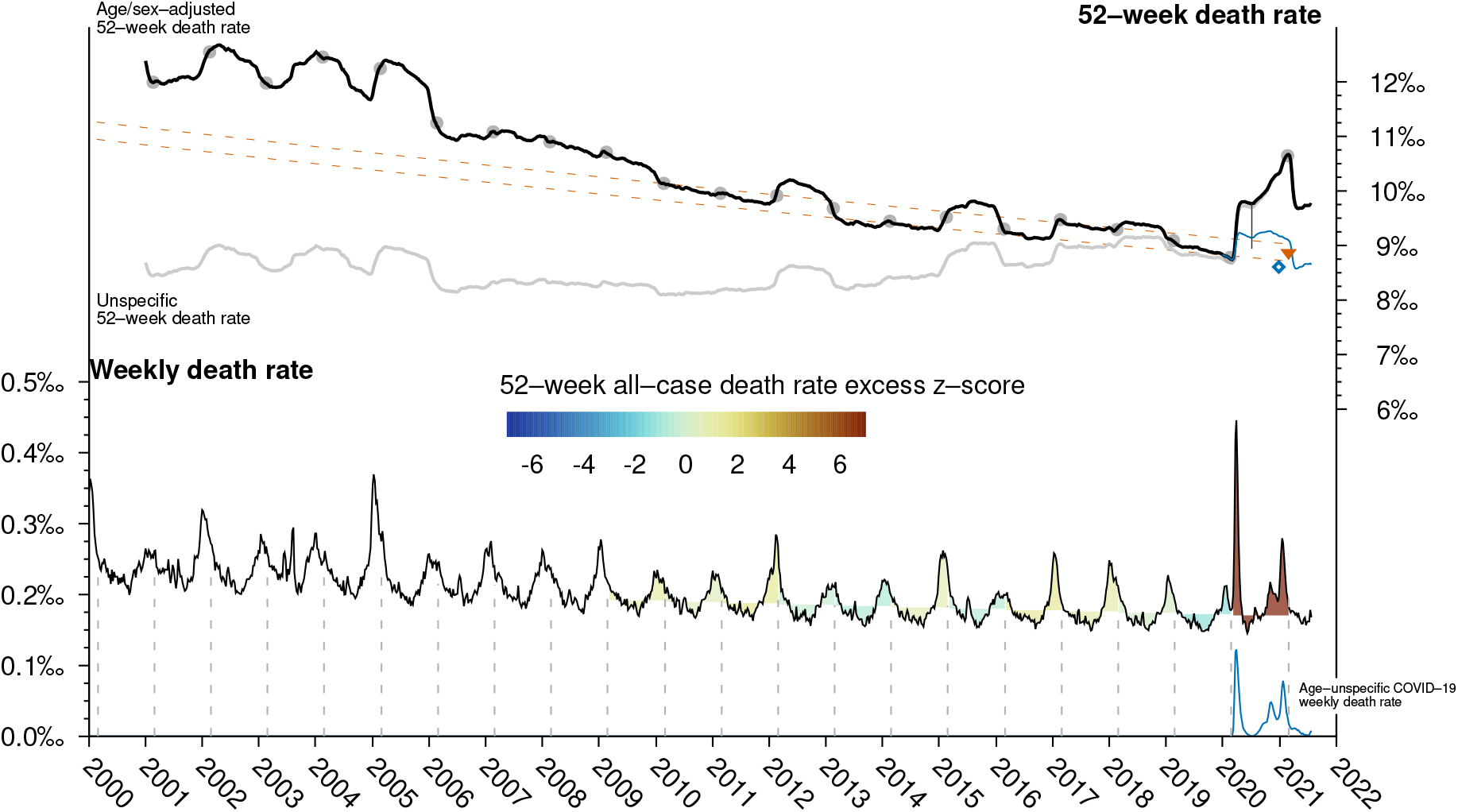
Bottom: all–cause, age/sex–adjusted weekly per capita death rate and COVID–19 weekly death rate (blue) in Spain since 2000. Top: cumulative 52–week, all–cause, age/sex–adjusted death rate (dark orange) and cumulative 52–week age/sex–adjusted, non COVID–19 death rate (blue). Dots display 52–week values at the week 08 of every year in the 21st century. The down triangle symbol shows the predicted 2021 death rate. The dashed lines around the 52–week death rate are centred at death rate predicted by the Poisson regression and is one unbiased sample standard deviation of the residuals prior to the pandemic onset in width. The vertical thin line notes the excess death rate at the end of week 27–2020. For the sake of comparison the unspecific, 52–week is shown in light grey. The 52–week cumulative death rate predicted by the Poisson model is converted into a weekly average to shade the weekly death rate accordingly. Net shaded area represents the 52–week accumulated death rate. Colour depth for shaded areas is given by the *z*−score. The diamond symbol signals the expected value deduced from Islam *et al*. [4] (see Discussion).

Circles in figure 2 display the observed values in week 08 for the years. Dashed lines display the predicted value of the Poisson regression plus and minus one unbiased sample standard deviation of the residuals (*s* = 157×10^−6^ or 7452 deaths if population numbers are used).

The blue line displays the age/sex–adjusted, non COVID–19 death rate which is obtained after removing the COVID–19 weekly deaths reported by the CNE, shown in the bottom of the figure. The triangle signals the expected, age/sex–adjusted, all–cause death rate for week 08–2021 which is 8.872 × 10^−3^, compared to the observed 10.662 × 10^−3^ rate.

Weekly deaths are shaded by the expected death rate obtained by regression. Therefore the net shaded areas display the net excess death rate. Colour depth is given by the normal score of the excess.

## 4. DISCUSSION

The goal of computing excess deaths is the goal of determining the predicted or reference value from which the excess is computed. To accomplish that, one requires on the first hand a record of previous observations to infer a prediction; the longer the record, the merrier. When it became evident that mortality record would be under close public scrutiny in 2020 Eurostat requested to national statistical offices to provide, if possible, weekly records of mortality starting in the year 2000.

On the other hand causality tends to challenge this idea: the older the observation the less impact may have in the future prediction. As an example Karlinsky and Kobak [5] considered data from the year 2015 onward in their world mortality data set while Islam *et al*. [4] considered data from the year 2016 onward on their analysis and deduced 84 100 excess deaths in the calendar year 2020. In this work the excess deaths on week 52–2020 is only 67 143.

There are two sources for the discrepancy. First since April 2021 INE recalculated 2020 weekly deaths and removed some 3500 deaths from their initial estimate, which would have yield a excess of ∼80 600 total excess deaths by Islam *et al*. [4] estimate. On the second hand, Islam *et al*. [4] considered only data from 2016 onward. Figure 2 shows in a diamond symbol the expected death rate that would have yield a total excess equal to 80 600 deaths at the end of the year 2020. It lies in the trend of past four years and on the lower bound of the trend observed in the past eleven years.

A important question to elucidate is the 52–week infection rate that produced the excess death rate. First column data in Table 2 lists the case rate —the ratio of the accumulated 52–week number of COVID–19 cases to population—. This metric is often challenged by the fact that due to a myriad of societal and health issues a fraction of cases often lies below the radar of epidemiological studies. In this pandemic this was specifically significant during the spring of the year 2020 due to limited testing capabilities.

**TABLE 2.**
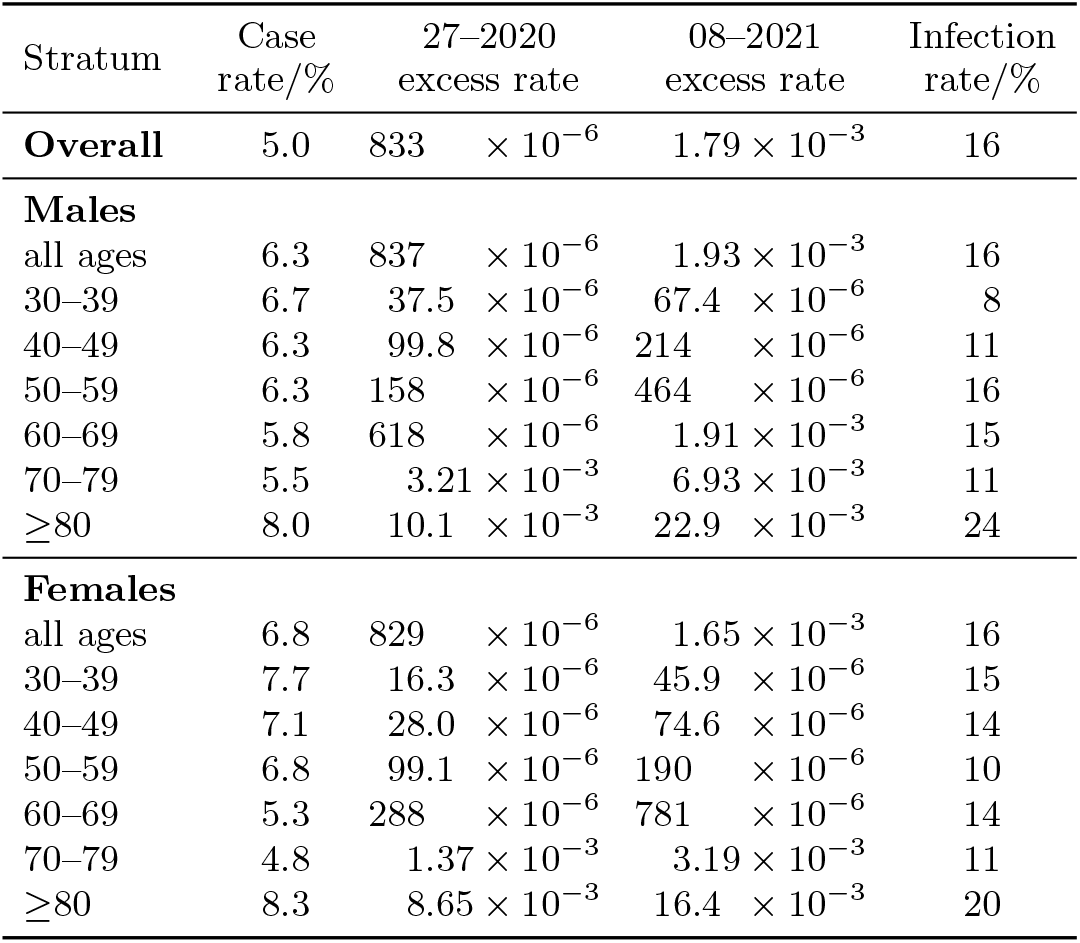
COVID–19 case and infection rates in Spain during the first year of the pandemic. The first data column displays the accumulated case rate as obtained from the CNE records. Second and third data columns list the excess death rate at week 27–2020 and at week 08–2021 from which the infection rate (fourth data column) is computed after removing 19 681 COVID–19 related deaths that occurred in institutionalised population.

| Stratum | Case<br>rate/% | 27–2020<br>excess rate |  | 08–2021<br>excess rate | Infection<br>rate/% |
| --- | --- | --- | --- | --- | --- |
| **Overall** | 5.0 | 833 | $\times 10^{-6}$ | $1.79 \times 10^{-3}$ | 16 |
| **Males** |  |  |  |  |  |
| all ages | 6.3 | 837 | $\times 10^{-6}$ | $1.93 \times 10^{-3}$ | 16 |
| 30–39 | 6.7 | 37.5 | $\times 10^{-6}$ | $67.4 \times 10^{-6}$ | 8 |
| 40–49 | 6.3 | 99.8 | $\times 10^{-6}$ | $214 \times 10^{-6}$ | 11 |
| 50–59 | 6.3 | 158 | $\times 10^{-6}$ | $464 \times 10^{-6}$ | 16 |
| 60–69 | 5.8 | 618 | $\times 10^{-6}$ | $1.91 \times 10^{-3}$ | 15 |
| 70–79 | 5.5 | $3.21 \times 10^{-3}$ | | $6.93 \times 10^{-3}$ | 11 |
| ≥80 | 8.0 | $10.1 \times 10^{-3}$ | | $22.9 \times 10^{-3}$ | 24 |
| **Females** |  |  |  |  |  |
| all ages | 6.8 | 829 | $\times 10^{-6}$ | $1.65 \times 10^{-3}$ | 16 |
| 30–39 | 7.7 | 16.3 | $\times 10^{-6}$ | $45.9 \times 10^{-6}$ | 15 |
| 40–49 | 7.1 | 28.0 | $\times 10^{-6}$ | $74.6 \times 10^{-6}$ | 14 |
| 50–59 | 6.8 | 99.1 | $\times 10^{-6}$ | $190 \times 10^{-6}$ | 10 |
| 60–69 | 5.3 | 288 | $\times 10^{-6}$ | $781 \times 10^{-6}$ | 14 |
| 70–79 | 4.8 | $1.37 \times 10^{-3}$ | | $3.19 \times 10^{-3}$ | 11 |
| ≥80 | 8.3 | $8.65 \times 10^{-3}$ | | $16.4 \times 10^{-3}$ | 20 |

To overcome this issue Pastor-Barriuso *et al*. [6] conducted during the spring of the year 2020 a seroepidemiological study (ENE–COVID) within community dwelling population which determined the SARS–CoV–2 seroprevalence or infection rate in Spain at the end on week 27–2020 *S*_27_ can be determined. If the infection fatality risk IFR —the ratio of (excess) deaths to infected population— is assumed to be invariant in time then the shares of infected population on week 8–2021 can be inferred from the proportional count *I*_08_ = *S*_27_×*E*_08_*/E*_27_. As noted by Pastor-Barriuso *et al*. [6] (supplementary method) a number of 19 681 COVID–19 related deaths that occurred within institutionalised population must be removed from *E*_27_ and *E*_08_ to assess the excess death rates within community dwelling population, which the seroprevalence refers to. The infection rate is listed in the last data column in Table 2. In many cases this metric climbs to three times the 52–week case rate reported from the CNE.

## 5. CONCLUSIONS

A grand total of 505 546 people died in Spain from March 2nd, 2020 to February 28th, 2021 the first year (52 weeks, 364 days) after the pandemic outbreak. This translates into a death rate equal to 10.662 × 10^−3^.

Age/sex–adjusted, 52–week death were built from age–specific (5–year bin) and sex–specific death rates.

Age/sex–adjusted, 52–week death rates had been smaller than 10.662 × 10^−3^ for the past continued 13 years.

A bivariate generalised Poisson regression for the last eight years of age/sex–adjusted, 52–week death rates predicted 8.872 × 10^−3^ for this period of time with an unbiased standard deviation of the residuals equal to 157 × 10^−6^ or 7452 deaths.

This results in an estimate excess death rate 1.790 × 10^−3^ with 95 % CI 1.773 × 10^−3^ to 1.808 × 10^−3^. This translates into 84 849 excess (84 008 to 85 690) death of which 73 571 deaths (87 %) were previously identified as COVID–19 deaths and the remaining 11 278 (13.3 %) were undercounts. Among them a number of 9772 COVID–19 related deaths (11.5 % of excess deaths) occurred in care facilities and nursing homes during the spring of the year 2020 (see Pastor-Barriuso *et al*. [6]). Therefore it is only 1496 deaths (1.8 % of excess deaths, a *z*−score of 0.2) that remain unattributed.

It is estimated that 16 % of the Spanish population was infected by the SARS–CoV–2 during the first year of the pandemic.

## Data Availability

Data used in this manuscript come from public records.

